# High-Dose Naloxone Formulations Are Not as Essential as We Thought

**DOI:** 10.1101/2023.08.07.23293781

**Authors:** Paige M. Lemen, Daniel P. Garrett, Erin Thompson, Megan Aho, Christina Vasquez, Ju Nyeong Park

**Affiliations:** Tennessee Harm Reduction, Jackson, TN, USA; University of Tennessee Health Science Center, Memphis, TN, USA; Harm Reduction Innovation Lab, Rhode Island Hospital, Providence, RI, USA; The Warren Alpert Medical School, Brown University, Providence, RI, USA

**Author notes:** Corresponding author: Paige M. Lemen Tennessee Harm Reduction 1989 Madison Avenue, 7, Memphis, TN 38104. Ethics approval and consent to participate: Not applicable. Consent for publication: Not applicable. **Availability of data and materials:** Data sharing is not applicable to this article as this is a literature review. However, a table (Supplementary Table 1) has been provided that lists all published research articles involved in the literature search for this review. The data referenced by Tennessee Harm Reduction are not publicly available because it was used only as observation. However, a summary of this data is available on the Tennessee Harm Reduction website and data from the corresponding author on reasonable request. **Competing interests:** The authors declare that they have no competing interests. **Funding:** MA is funded by the Karen T. Romer Undergraduate Teaching and Research Award at Brown University. ET and JNP are funded by the Center of Biomedical Research Excellence on Opioids and Overdose (P20GM125507) from the NIH. JNP serves as a technical consultant for a modeling project funded by the Food and Drug Administration (U01FD00745501) based at Harvard Medical School. Its contents are solely the responsibility of the authors and do not necessarily represent the official views of our funders. **Author Contributions:** PML and DPG conceptualized and drafted the first version of the paper. All authors contributed to writing subsequent versions and have approved the final manuscript.

**Keywords:** naloxone, opioid overdose, literature review

## Abstract

Naloxone is a U.S. Food and Drug Administration (FDA) approved opioid antagonist for reversing opioid overdoses. Naloxone is available to the public, and can be administered through intramuscular (IM), intravenous (IV), and intranasal spray (IN) routes. Our literature review aimed to improve understanding regarding the adequacy of the regularly distributed two doses of low-dose IM or IN naloxone in effectively reversing fentanyl overdoses and whether high-dose naloxone formulations (HDNF) formulations are an optimal solution to this problem. Moreover, our initiative incorporated the perspectives and experiences of people who use drugs (PWUD), enabling a more practical and contextually-grounded analysis. We began by discussing the knowledge and perspectives of Tennessee Harm Reduction, a small peer-led harm reduction organization. A comprehensive literature review was then conducted to gather relevant scholarly works on the subject matter. The evidence indicates that, although higher doses of naloxone have been administered in both clinical and community settings, the vast majority of fentanyl overdoses can be successfully reversed using standard IM dosages with the exception of carfentanil overdoses and other more potent fentanyl analogs, which necessitate three or more doses for effective reversal. Multiple studies documented the risk of precipitated withdrawal using high doses of naloxone. Notably, the possibility of recurring overdose symptoms after resuscitation exists, contingent upon the half-life of the specific opioid. Considering these findings and the current community practice of distributing multiple doses, we recommend providing at least four standard doses of IN or IM naloxone to each potential bystander, and training them to continue administration until the recipient achieves stability, ensuring appropriate intervals between each dose. Based on the evidence, we do not recommend HDNF in the place of providing four doses of standard naloxone due to the higher cost, risk of precipitated withdrawal and limited evidence compared to standard IN and IM. All results must be taken into consideration with the inclusion of the lived experiences, individual requirements, and consent of PWUD as crucial factors. It is imperative to refrain from formulating decisions concerning PWUD in their absence, as their participation and voices should be integral to the decision-making process.

## INTRODUCTION

Naloxone hydrochloride is a life-saving opioid antagonist medication that can reverse the effects of opioid overdose [1]. Naloxone has been shown to be one of the most effective interventions in reducing opioid-related deaths at the population level [2]. Opioid receptor antagonists in the central or peripheral nervous system inhibit one or more opioid receptors[3,4]. This means that it counteracts both the expected (such as analgesia and euphoria) and unintended (such as respiratory depression and coma) effects of opioid agonists, the latter of which is ultimately what causes death from opioid overdose. Naloxone administered by community members (people who use drugs [PWUD], friends, family, etc) has proven successful in reversing opioid overdoses in 75-100% of cases[5]. It is considered safe at the recommended doses to opioid-naive persons as well. Naloxone is primarily available to the public through free community-based programs though some naloxone is accessed through provider prescriptions, and pharmacy-initiated distribution (e.g., through state-level standing order policies).

There are three United States Food and Drug Administration (FDA)-approved administrations of naloxone that are available to PWUD: injectable intramuscular (IM), intravenous (IV), and intranasal spray (IN); subcutaneous auto-injectors have been discontinued [6][7]. These formulations as well as their brand names are described in Table 1. One of the most popular formulations of naloxone supplied to PWUD in harm reduction programs is generic (unbranded) IM naloxone kits[8,9]. Injectable naloxone solution is typically provided in two or three vials in IM naloxone kits[5]. 1-mL vials with 0.4 mg/mL are the only available form. 10-mL vials containing 0.4 mg/mL are also available but are not readily accessible in IM naloxone kits provided to PWUD[10,11]. Regardless of the exact route of administration, naloxone can be easily administered by a lay bystander. After naloxone administration, the person should be monitored for signs of recovery[16].

The FDA has continued to support high-dose naloxone formulations (HDNF) such as Kloxxado (double the dosage of standard IN) and Zimhi (25 times higher dose than generic IM)[17], despite the voiced concerns of harm reduction workers and others with real-world experience of regularly using naloxone to reverse and overdose[12,13]. This is in part due to concerns that the increased potency of fentanyl and fentanyl analogs compared to heroin and prescription opioids could require higher doses of naloxone to be effective. Having input on drug policy and research from people with lived experiences of drug use is important and useful because it allows for a more comprehensive understanding of the complexities and realities of drug use, ensuring that policies and research are informed by the perspectives and needs of those directly affected, leading to more effective and equitable outcomes[14]. However, the historical exclusion of these individuals such decisions can be attributed, in part, to the pervasive stigma surrounding drugs, people who use them, and those with substance use disorders (SUD)[15].

It’s important to note that drug use is a complex issue that is influenced by a variety of factors, such as social, economic, environmental, and other social determinants of health. Moreover, individuals engaged in drug use should not be delineated solely by their drug consumption, but rather recognized as multifaceted individuals with distinct requirements and aspirations. Therefore, adopting an impartial and non-judgmental approach is imperative when addressing the subject of drug use and, consequently, the application of naloxone.

The criminalization of drug use has resulted in stigma and fear among the general population which, in turn, has led to a misunderstanding of and opposition towards harm reduction strategies. In the context of naloxone, the misunderstandings can be seen in the altered portrayal of the effects and risks associated with naloxone use for overdose reversal. For example, stigma can lead to new developments regarding appropriate naloxone dose and administration route without input from people with living experience resulting in products that the at-risk population does not need or want.

Opioid dependency necessitates individuals to consume opioids once or more daily, regardless of expressed concern around naloxone misuse and overuse[[3,16]. This is particularly a concern for those who use shorter-acting opioids such as fentanyl and its analogs[17], which presently dominate the US illicit market for opioids[17,18]. Moreover, the heightened potency of fentanyl, fentanyl analogs, coupled with the absence of quality controls in illicit markets and non-pharmaceutical-grade manufacturing processes, leaves opioid-dependent individuals particularly vulnerable to overdose through misjudging their dosage[19]. This concern for high overdose rates and drug poisoning severity provides theoretical justification for the proposal of higher doses of naloxone formulations. Nevertheless, this theoretical foundation lacks input from the ground level experts: PWUD, harm reduction workers, and other relevant groups possessing firsthand experience and direct involvement with drug users.

Accordingly, the goal of this literature review is to improve understanding on how often more than two doses of IM and IN naloxone are needed in the community to reverse a fentanyl overdose and whether promoting HDNF is an optimal and necessary solution to this problem. There were three literature reviews conducted on this topic previously: Moe and colleagues conducted a A rigorous systematic review of overdoses (n=26,660) from North America and Europe through 2018 and found that although higher initial and cumulative naloxone doses were being used by lay and healthcare responders for overdoses presumed to be fentanyl or another synthetic opioid, a cumulative total dose of 4mg of naloxone (e.g., two doses of standard IN) was sufficient in 97% of presumed fentanyl/potent opioid cases [20]. Abdelal and colleagues (2022) recently examined the use of two or more naloxone doses, however the authors received consultancy fees or stock options from Hika Pharmaceuticals, which manufactures the HDNF product Kloxxado. Chou and colleagues (2017) only compared standard IN vs. IM doses. The response to opioid overdose with naloxone remains an imprecise science despite its reliability in reversing opioid overdose and having negligible contraindications for those who are not physiologically dependent on opioids.

## Background

### I. Responding to an Overdose

In the case of a suspected opioid overdose, a five-step process is recommended for bystanders to provide effective and timely care[21]. The first step involves checking for signs of opioid overdose, such as unconsciousness, slow or absent breathing, pale and clammy skin, and slow or no heartbeat. Next, the bystander immediately calls emergency medical services (EMS) to ensure timely medical attention. They then administer naloxone and clear the airways to perform rescue breathing to help provide oxygen to the body until medical help arrives. The goal of naloxone is to help the person breathe normally again, but not necessarily complete alertness[22]. The current recommendation by the Substance Abuse and Mental Health Services Administration (SAMHSA) is to administer another dose of naloxone if the person overdosing does not show improvement within 2-3 minutes, and to repeat this step using that time frame[19]. Finally, the bystander should stay with the person and monitor their response until EMS takes over care. Following this recommended five-step process can greatly improve the chances of a positive outcome for the patient experiencing an opioid overdose[21].

Administering oxygen to a person experiencing an opioid overdose is a recommended additional step in the resuscitation process[23–25]. Supplemental oxygen helps ensure the individual receives adequate oxygenation, which is essential for sustaining vital organ function. However, it is important to note that oxygen administration alone is not considered a definitive treatment for an opioid overdose. Oxygen equipment or someone cardiopulmonary resuscitation (CPR) certified is also not often available. Naloxone administration, along with providing oxygen when possible, can greatly enhance the chances of successful resuscitation and recovery[24]. Therefore, it is advisable to promptly administer naloxone in addition to oxygen when dealing with suspected opioid overdoses.

### II. Naloxone options in the U.S

The U.S. currently has seven overdose reversal products that contain naloxone described both below and in **Table 1**.

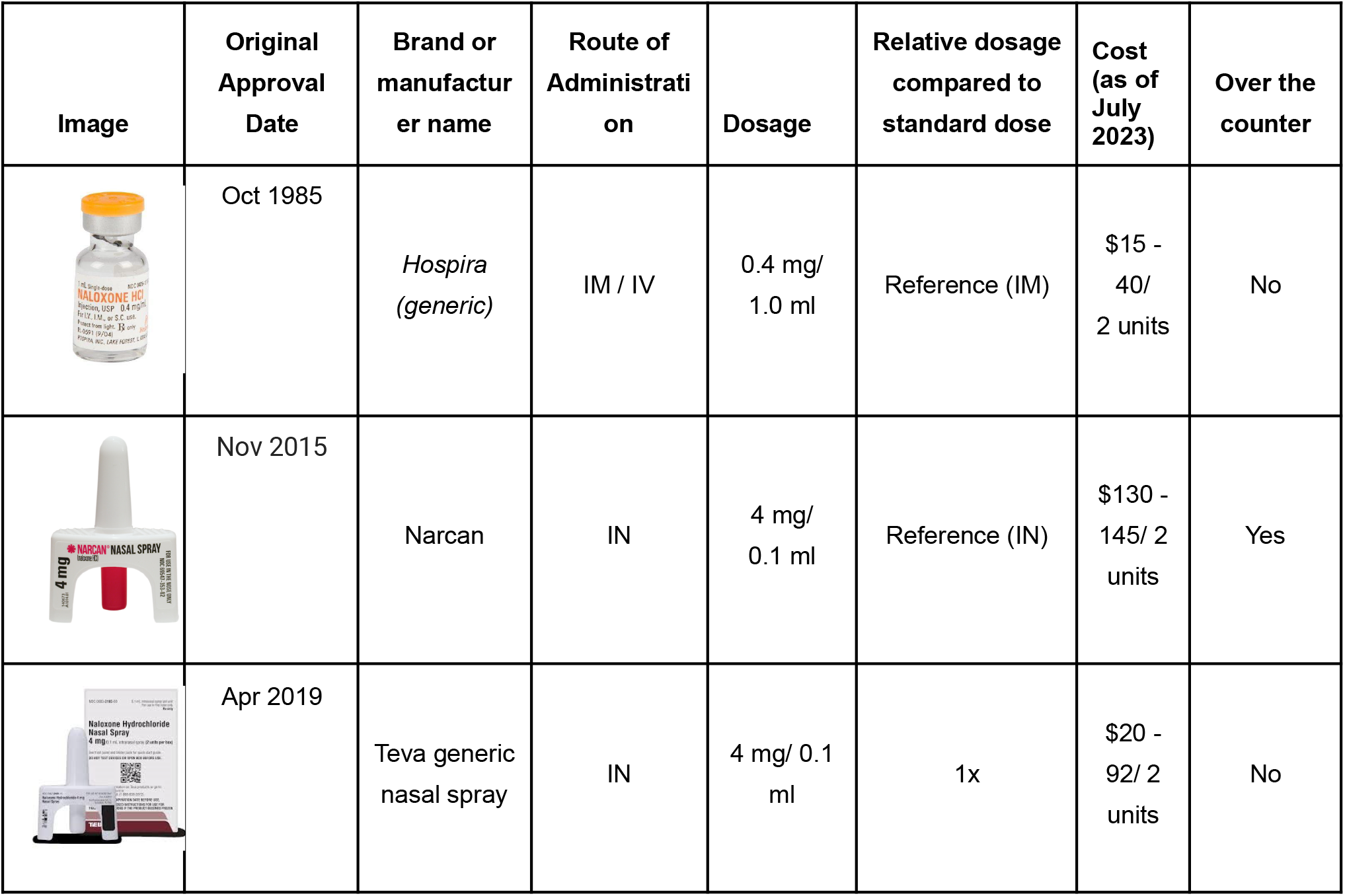
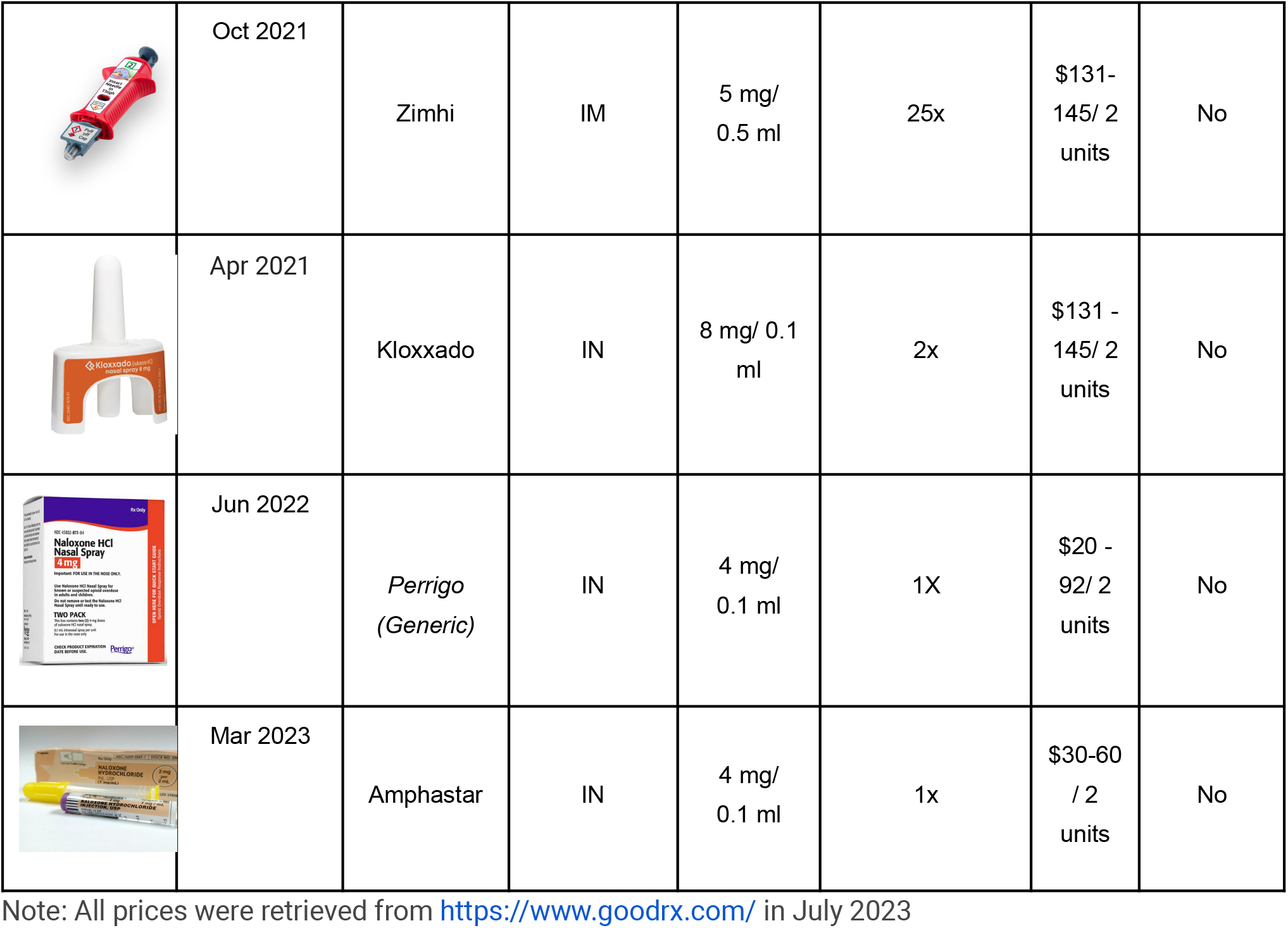

| Image | Original Approval Date | Brand or manufacturer name | Route of Administration | Dosage | Relative dosage compared to standard dose | Cost (as of July 2023) | Over the counter |
| --- | --- | --- | --- | --- | --- | --- | --- |
| | Oct 1985 | Hospira (generic) | IM / IV | 0.4 mg/ 1.0 ml | Reference (IM) | \$15 - 40/ 2 units | No |
| | Nov 2015 | Narcan | IN | 4 mg/ 0.1 ml | Reference (IN) | \$130 - 145/ 2 units | Yes |
| | Apr 2019 | Teva generic nasal spray | IN | 4 mg/ 0.1 ml | 1x | \$20 - 92/ 2 units | No |
| | Oct 2021 | Zimhi | IM | 5 mg/<br>0.5 ml | 25x | \$131-145/ 2<br>units | No |
| | Apr 2021 | Kloxxado | IN | 8 mg/ 0.1<br>ml | 2x | \$131 -<br>145/ 2<br>units | No |
| | Jun 2022 | Perrigo<br>(Generic) | IN | 4 mg/<br>0.1 ml | 1X | \$20 -<br>92/ 2<br>units | No |
| | Mar 2023 | Amphastar | IN | 4 mg/<br>0.1 ml | 1x | \$30-60<br>/ 2<br>units | No |

Note: All prices were retrieved from https://www.goodrx.com/ in July 2023

Generic injectable naloxone comes in 1mL vials of 0.4mg/mL concentration[5]. It can be utilized in any method of administration, including intramuscular (IM), and intravenous (IV) routes, as well as intranasally (IN) via an atomizer. It can also be administered subcutaneously (SC),although it is not marketed for this route. NARCAN^®^ Is the most well-known brand name for naloxone nasal spray. It comes with a 0.1 mL pre-packaged solution that contains 4mg/0.1mL of naloxone. This specific brand’s pre-packaged administration tool only allows for naloxone to be administered intranasally. The generic counterparts to NARCAN^®^ are the Teva and Perrigo generic nasal sprays which have chemically identical active ingredients and concentrations. Kloxxado^®^ is a newer naloxone nasal spray that also comes with double the dose of NARCAN. It comes with a 0.1 mL pre-packaged solution that contains 8 mg/0.1mL of naloxone. Identical to NARCAN, this specific brand’s pre-packaged administration tool also only allows for naloxone to be administered intranasally. Zimhi is a brand name syringe that comes with 0.5 mL of 5mg/0.5 mL naloxone. This syringe can only be administered intramuscularly (IM). Lastly, Amphastar^®^ Prefilled Naloxone Syringes come in 2 mL, with 1mg/mL naloxone. These prefilled syringes are also compatible with any method of administration, both nasal and injection routes. It is a recently approved IN naloxone formulation with a 4 mg/0.1mL concentration. It is not yet widely available in the US.

In May 2023, the FDA approved Opvee®, a nasal spray version of nalmefene and the first alternative opioid antagonist indicated for opioid overdose reversal. The half-life of nalmefene is ∼11 hours, much longer than the 60- to 90-minute half-life of naloxone[26]. Nalmefene may reverse opioid intoxication for longer than naloxone, which some view as a benefit over naloxone. However, its extended half-life presents the continued concern of placing opioid-dependent persons who overdose into precipitated opioid withdrawal for far longer than naloxone. Opvee was developed by Opiant Pharmaceuticals, which plans to release Opvee to the U.S. market as early as October 2023[27]. Opiant also developed Narcan. However, while Opiant manufactures Narcan, Emergent Operations Ireland LTD produces and holds the FDA approval.

### IV. Bioavailability

Bioavailability, referring to the proportion of a drug that is able to enter the body’s circulation to have an active effect[28] and is a key consideration to take into account during discussions of naloxone dosing. It is used to approximate a drug’s effectiveness when taken by a patient. Factors that can affect bioavailability include the administration route (e.g., oral, intravenous, intramuscular), drug form (e.g., capsule, tablet, liquid), and personal health history (e.g., age, weight, liver function).

The IN and IM administration routes are both commonly used by first responders in the community. They have bioavailability of 50% and 98% and a latency time to effectiveness of 15-minutes and 8-minutes, respectively[26,28–30]. The IV route is only used in inpatient medical settings and has a bioavailability of 100% with a 2-minute latency to effect. The IV route is preferred by medical professionals as the dose can be tailored to each patient allowing for sufficient overdose reversal while minimizing the risk of withdrawal.

### V. The Consequences of Preconceived Biases towards PWUD

There is a considerable amount of opposition towards drug use and harm reduction for a variety of reasons, some of which include a lack of understanding, stigma, fear, political beliefs, personal experiences, misinformation and criminalization[31–33]. The prevailing societal perceptions, dissemination of misleading information through previous propaganda, and the influence of the “war on drugs” have contributed to the pervasive negative attitudes towards drug use held by a substantial portion of the population. Now, due to the criminalization of many drugs, individuals who use recreationally for personal, constructive purposes, and in a manner characterized by safety and responsibility, remain largely invisible within media representation and public forums[34]. Instead, those who suffer from a SUD, overdose, or those only in abstinence-based recovery are displayed. This biased depiction results in an altered portrayal of the effects and risks associated with recreational drug use[35] and provides opportunity for reinforcing racial, gender, and class stereotypes pertaining to drug users[35,36]. Additionally, this also means stigma can contribute to more unnecessary challenges to deciding on appropriate naloxone dose and administration routes.

## Methods

Initially, we introduced and assessed the knowledge and perspectives contributed by Tennessee Harm Reduction, a drug-user-run community-based organization in rural West Tennessee focused on distributing naloxone, drug checking supplies, and safer use supplies to over 200 community members. We specifically drew from the valuable expertise of two experienced harm reductionists: Daniel Garrett (DPG), the founding director, and Paige Lemen (PML), a Research and Outreach Specialist. Through this work, DPG and PLM made several noteworthy findings including that IN naloxone was reported by the community to cause worse precipitated withdrawal when compared to IM naloxone and seemed to take longer to revive the overdosing individual. As a result, people giving or receiving naloxone became cautious about continuing to use it. Tennessee Harm Reduction recorded ∼215 known overdose reversals (not including reversals performed on pets) among those who utilize their services. ∼135 reversals were known to have given IM naloxone first, and about half of those only needed one dose. A further A further ∼40% required two doses with the remaining ∼10% having 3 or more doses administered. No unsuccessful overdose reversals have been reported. It is important to note that both time between doses and whether the administrator had proper training was not reported so it is unclear whether 3 or more doses were actually needed. HDNF caused even worse symptoms of precipitated withdrawal. DPG also noted that newly available HDNF cost between 2 to 10 times more than generic IM naloxone.

As one of few harm reduction programs in Tennessee that primarily distributes IM naloxone, we saw a need to better understand the evidence behind whether adopting newer HDNF would be beneficial. Through an unfunded partnership with Harm Reduction Innovation Lab, a search strategy was developed and implemented between August 2022 and February 2023. Phrases included: “high-dose naloxone formulation” and “opioid overdose.” Other keywords searched were “naloxone dosage overdose”, “naloxone dosage”, “high dose naloxone”, “high dose naloxone opioid”, “naloxone dosing”, “naloxone formulation”, “high dose naloxone formulation”, “high-dose naloxone”, and “high-dosage naloxone”. An exhaustive literature review was performed by applying this strategy to search engines such as PubMed, Google,and Google Scholar to compile a comprehensive collection of relevant scholarly works. We filtered for original peer-reviewed articles published between January 2012 to February 2023. We also conducted a Google search to gather information on each naloxone product and cited research. We reviewed the title and abstract of each article and excluded those that focused on unrelated concepts including buprenorphine, specific health diagnoses and disorders, or animal and computer models.

The remaining eligible articles were downloaded, read and summarized by a research assistant (MA) using a synthesis matrix developed by the study team. The matrix was designed to extract article characteristics, including whether the overall sentiment supported or opposed HDNF, any funding received, the authors’ employment, any conflicts of interest, overall summary of the concepts researched, key conclusions, the frequency two doses of naloxone were needed or administered, and the stated advantages and disadvantages of HDNF (if applicable). Finally, the findings were discussed by the full team and organized into three topics as described below. Given the number of articles funded by naloxone manufacturers or consultants paid by pharmaceutical companies, we decided to flag those articles and discuss them in a separate section in order to minimize bias.

## Results

We successfully identified 22 articles eligible for inclusion (**Figure 1**). Seven articles (32%) were coded as supportive of HDNF and four (18%) were coded as unsupportive of HDNF with the remainder being neutral with arguments for and against HDNF (**Table 2**). Notably, very few articles elicited the perspectives of PWUD. The main findings of our synthesis are organized under three overarching questions: (1) how often are more than two standard doses of naloxone needed to reverse a fentanyl overdose; (2) what is the evidence for HDNF; and (3) what is the evidence against HDNF.

**Figure 1.**
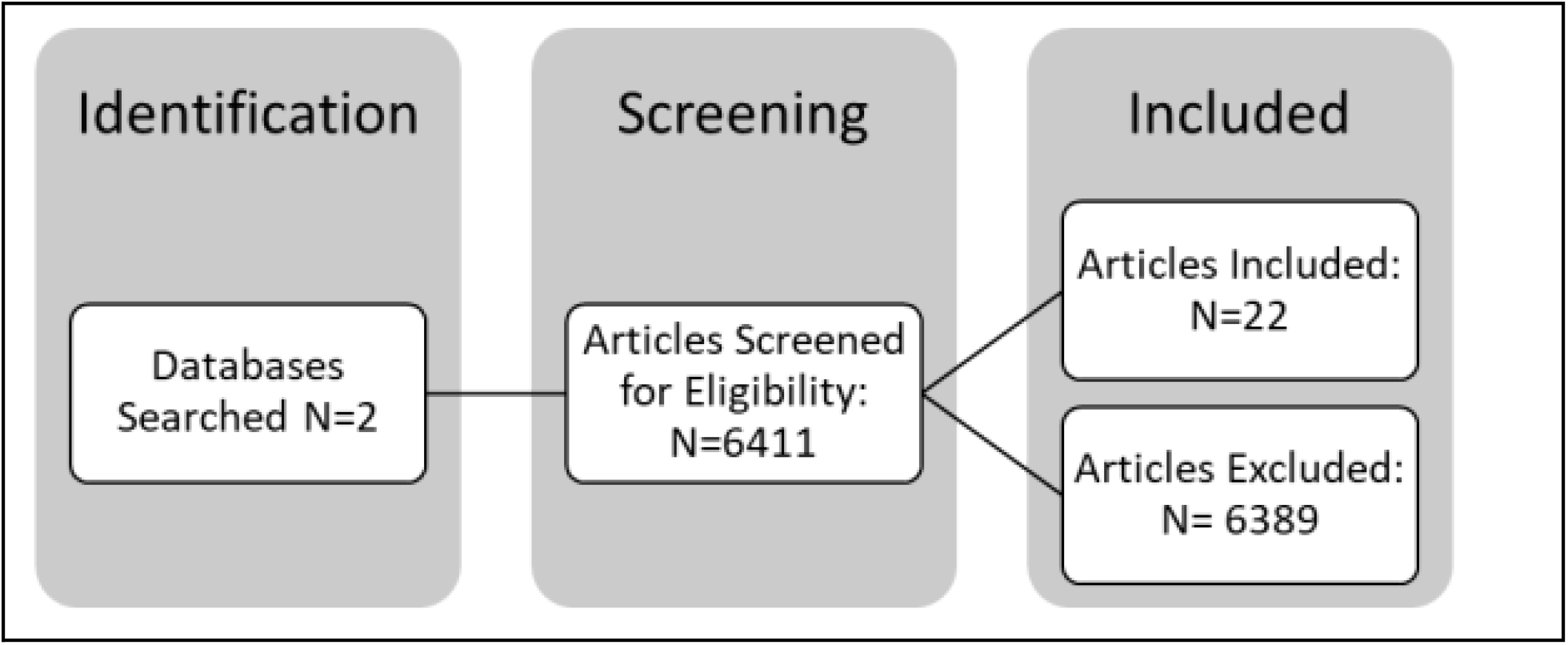
Flow chart of all articles included

### PART ONE: HOW OFTEN ARE MORE THAN TWO STANDARD DOSES OF IM or IN NALOXONE NEEDED TO REVERSE A FENTANYL OVERDOSE?

Several factors determine whether multiple doses of naloxone are needed to reverse an overdose. These factors include the type, amount and method of opioid use, individual opioid tolerance levels, the individual overall health status, and naloxone administration route. Some people may require multiple doses of naloxone either immediately or as the effects of naloxone may wear off before those of longer lasting opioids[37].

We found relatively few published papers examining the number of doses of IM or IN naloxone needed to reverse an overdose. Supporters of HDNF often reference a single nationwide study conducted from 2012-2015[38], which reported that Emergency Medical Service (EMS) providers needed to administer multiple doses of naloxone, with increases each year. However, this study is limited in its methodology as it fails to provide detailed information regarding the administration method of naloxone utilized. This lack of transparency in the study design may impede the generalizability of the findings. Other researchers report that there was an increase in the average dose provided by EMS; from 2014 and 2016, in Ohio the average IV dose increased from 2mg to 5mg[39].

A study conducted in Pittsburgh from 2013-2016 found that less than 5% of overdoses required three or more doses of IM naloxone[40]. This was corroborated by a national study though heroin was involved in the majority of these cases[41]. Only two percent of EMS responders in New Jersey reported requiring a third dose of IN naloxone after having a second dose administered by an advanced life support team [42]. A large study of New York police officers from 2015-2020 noted that an average of two doses of naloxone were administered to rescue individuals[43]. One survey-based study reported that 30% of participants living in regions with fentanyl epidemics used 3 or more doses of IN naloxone[37].

Interestingly, a Morbidity and Mortality Weekly Report (MMWR) reported 83% of patients in Massachusetts required 3 or more doses of nasal naloxone to reverse a suspected fentanyl overdose[44]. A 2021 survey-based study out of Maryland indicated that 79% of participants administered 3 or more doses at their last witnessed overdose[45]. Case studies have recorded high doses (12-15 mg) of naloxone being administered for synthetic opioid overdoses[46]. More than two doses of naloxone were required to reverse two carfentanil overdoses, likely owing to the greater affinity of carfentanil for μ-receptors than naloxone[47]. μ-opioid receptors are one of the specific target sites in the body that naloxone, a medication used for opioid overdose reversal, binds to and blocks, effectively reversing the effects of opioid drugs[48,49].

An aforementioned rigorous systematic review by Moe and colleagues of overdoses (n=26,660) from North America and Europe through 2018 found that less patients with presumed fentanyl/ultra-potent opioid exposure were revived using initial low doses (≤0.4mg/ml) versus when heroin was presumed (57% vs. 80%) but they concluded that a cumulative dose of 4mg (e.g., two standard doses of IN) was sufficient in 97% of presumed fentanyl/potent opioid cases [20].

In conclusion, although there have been greater doses used in clinical and community settings, evidence suggests that the vast majority of fentanyl overdoses can be reversed with standard dosing. However, overdoses involving carfentanil and other potent synthetic analogs may require three or more doses. Two doses of IM naloxone (0.8mg) has also been insufficient in reversing some fentanyl overdoses though such data are subject to the amount of fentanyl actually present in drug samples[50]. Depending on the half life of the opioid used, an individual may fall back into an overdose after being revived due to naloxone’s half life of 30-90 minutes[51]. Given these facts, and the observation that three or more doses are already being used in the community, our recommendation is that four doses of IN or IM naloxone be provided to community members with clear education on the length of latency to effectiveness to optimize coverage. To determine whether HDNF is beneficial we weigh the advantages and disadvantages of these formulations next.

### PART TWO: WHAT ARE THE POTENTIAL ADVANTAGES OF HIGH-DOSE FORMULATIONS?

There are many perceived benefits of HDNF, though, in practice, the evidence base is underdeveloped. Given that the formulations currently approved have been relatively comparable in their concentration to others of the same administration route, much of the literature has focused on administering multiple doses over time (“titration”) rather than administering a single HDNF. According to the small number of papers[29,52] on this topic, a HDNF could result in a faster response and reduce the magnitude of the harmful non-fatal impacts of drug toxicity, including cognitive and physiologic issues. A HDNF would improve reversal rates for overdoses involving carfentanil and other opioids that have a stronger μ-receptor affinity than fentanyl[47], though such experiences are relatively rare and localized. A recent national study showed that almost half (48%) of people who had reversed an overdose with naloxone held no preference for or against HDNF while over one third (36%) preferred a HDNF to be made available[13].

Owing to the preconceived biases around drug use and naloxone by association, the need to carry fewer doses may help reduce experiences of stigma[37], while simultaneously providing more convenience in portability. It is clear that one or two doses of standard IM and IN will not be sufficient to reverse all opioid overdoses. Continuing to administer supposedly suboptimal doses that fail to reverse overdoses may reduce the acceptance of naloxone in the community. Increasing options for consumers to choose from by complementing existing formulations could increase accessibility and decrease costs through market competition. Despite the benefits and community interest in HDNF,[13]see many potential pitfalls of relying solely on them.

### PART THREE: WHAT ARE THE POTENTIAL DISADVANTAGES OF HIGH DOSE FORMULATIONS?

As discussed previously, prior literature suggests that, despite fentanyl poisoning becoming more prevalent, a standard dose can still be equally effective in many cases[12,40,53–55]. Below, we discuss further reasoning to not recommend HDNF over standard dosing.

#### I. There is no pharmacological basis for high dose naloxone when it comes to fentanyl

In vivo, in the human brain, researchers have used a positron emission tomography (PET) scanner’s non-tomography positron detecting system to measure the dose-response curve of naloxone and found that ∼13μg/kg (0.013mg/kg) of naloxone per kg of patient bodyweight was required to produce an estimated 50% receptor occupation when given intravenously[56]. In general, a drug typically needs to occupy a sufficient number of its target receptors to initiate the desired biological response. Studies have indicated that achieving approximately 50% receptor occupation by naloxone is associated with its desired therapeutic effects in reversing opioid overdose[56,57]. This suggests that higher doses of naloxone may not be needed as long as 50% of the μ-opioid receptors are occupied. However, there are many factors that could affect level of occupancy such as route of administration and bioavailability[58].

A second pharmacokinetic consideration is the variable binding affinities of various opioids relative to the antagonistic effects of naloxone. Each opioid has a unique binding affinity (K_i_) towards the μ-opioid receptors. Naloxone must have a lower K_i_, indicating a stronger binding affinity, to successfully reverse an overdose. Notably, morphine and fentanyl have similar K_i_ values despite their vastly different potency levels demonstrating that potency does not correlate with binding affinity[59,60]. Therefore, stronger analogs are not an indication of the need for HDNF in the absence of binding affinity assessment.

While naloxone exhibits a relatively rapid and strong binding affinity to opioid receptors, it has a relatively short duration of action (60-90 minutes), meaning it is relatively quick to dissociate from the receptors[61,62][57], [61,62]. For longer acting opioids, this means opioids may rebind causing a recurrence of respiratory depression. HDNF has been shown to extend the duration of action but not the initial time between administration and overdose reversal[63]. The duration of a drug’s effects can vary depending on several factors, including the individual’s tolerance, the method of administration, the dosage, and the purity of the drug. HDNF may have an application for these longer-acting opioids and circumstances, but evidence is limited.

#### II. The Risk of Withdrawal From High Dose Naloxone

As with any medication, there are potential risks associated with taking too much naloxone. The main risk of excessive naloxone dosing is that it can cause rapid-onset naloxone-induced withdrawal symptoms if a person has a high dependence to opioids[64–66]. Naloxone works by binding to the same brain receptors as opioids at much higher affinity. It is effective at reversing overdose as it displaces opioids from the receptors without activating their sedative and respiratory depressant effects. The displacement effectively reverses the effects of the opioids and causes withdrawal even if opioids remain in the person’s system[67,68]. This can include the well-known symptoms such as severe pain, agitation, muscle cramps, and nausea[69]. Additionally, precipitated withdrawal has serious symptoms, such as diarrhea, vomiting, myalgia, anxiety, and autonomic hyperactivity[70]. Additionally, in rare cases, naloxone can cause an allergic reaction, such as hives, difficulty breathing, or swelling of the face, lips, tongue, or throat[71]. Sequelae such as death, coma, and encephalopathy have been documented in association with these occurrences. Notably, such events have predominantly manifested in patients with pre-existing cardiovascular disorders or those concurrently administered medications with comparable adverse cardiovascular effects. However, establishing a definitive cause-and-effect relationship requires further investigation[72].

Due to these risks, the recommended dose for opioid reversal remains controversial. The aforementioned withdrawal risk can lead to hesitation among PWUD when encountering a potential overdose. They must quickly balance the potentially life threatening consequence of withholding the narcan with the ensuing implications of withdrawal. Namely, that the intense discomfort and cravings following withdrawal can lead to subsequent increased use and opioid seeking behaviors. Additionally, negative experiences related to overdose reversal may result in avoidance of treatment due to fears of having similar experiences in an already stressful medical setting. Finally, withdrawal can impair an individual’s ability to carry out acts of daily living such as caring for oneself, attending work, or engaging in social activities. HDNF are likely to intensify these drawbacks as administration to someone with high opioid dependence can lead to more intensified symptoms then the typical dose would as more opioids will be displaced with naloxone.

#### III. The need for respect, consent, and a voice in drug policy: Ethical considerations

From a literal perspective, consent is the act of voluntarily agreeing to participate in something, such as a medical procedure, sexual activity, or research study. It is important because it ensures that individuals know and understand what they are agreeing to. Consent is a fundamental aspect of respecting individual autonomy and personal freedom and it is crucial for maintaining ethical standards in healthcare, research and interpersonal relationships.

In the context of an overdose, obtaining consent is not possible because they are unconscious. Unless the responder and person experiencing the overdose had discussed their preferences on how to handle such a situation before the overdose occurred, standard guidance should aim to do as little harm as possible. In such cases, it’s also important to provide clear and accurate information and to respect the autonomy of the person experiencing the overdose as much as possible after administration of naloxone when conscious.

Listening to PWUD is crucial in making informed decisions about increasing the dose formulation of naloxone. Those with lived experience have unique insight into the complexities of overdose and the effectiveness of naloxone. They can provide valuable information on how a higher dose formulation may impact their ability to respond to an overdose. Additionally, they can offer insight into other factors that may contribute to overdose, such as polysubstance use or lack of access to harm reduction services. By listening to those with lived experience, we can gain a better understanding of the challenges and barriers faced by PWUD and make more informed decisions about how to address overdose in a way that is effective, equitable, and inclusive.

Collaborating with PWUD is also an important aspect of practicing informed consent. By actively hearing their experiences and concerns, we can better understand their needs and preferences allowing us to provide care with respect and consideration of their unique circumstances. Individuals who use drugs have the right to make informed decisions about their healthcare and incorporating their preferences ensures they are empowered to make informed decisions about their healthcare. This can help build trust between healthcare providers and PWUD, leading to better health outcomes and more effective overdose prevention strategies. Conrarily, making decisions on naloxone dose, route of administration, and cost without including those who are directly impacted in the decision process violates their right to consent, erodes their trust and perpetuates the overdose epidemic.

#### IV. Cost considerations

As shown in **Table 1**, the costs of available naloxone formulations vary widely from $15-$40 per unit for the most affordable generic IM formulation to $131-$145 per unit for Zimhi high-dose IM auto injector and Kloxxado high-dose IN. As expected, generic formulations cost less than branded formulations with the IM and IN costing $15 and $20 at the lower cost range respectively. Notably, the two highest single dose formulations, Zimhi and Kloxxado, are also the most costly. Zimhi is 25 times stronger than generic IM naloxone and costs over 8 times the generic equivalent. Kloxxado is twice as strong as the generic IN formulation and costs about 5 times as much. Given that the majority of fentanyl overdoses studied only require two or three doses of standard IM or IN, high dose naloxone formulations with more than three times the dose within the same administration route category may not be a cost effective solution.

## DISCUSSION

Our review aimed to improve understanding regarding the adequacy of two doses of IM or IN naloxone in effectively reversing fentanyl overdoses in the community as an avenue to determine whether higher-dosage formulations are an optimal and necessary solution to this problem. Our findings indicate that, although two or more standard doses of naloxone have been administered in both clinical and community settings, the majority of fentanyl overdoses can be successfully reversed using three standard dosages of IN or IM. Overdoses involving carfentanil and other potent synthetic fentanyl analogs necessitate three or more doses for effective reversal; this may be due to carfentanil having a slower rate of opioid receptor dissociation [73]. However, carfentanil overdoses are relatively rare compared to fentanyl overdoses throughout the United States.

Although comparing formulations was beyond the scope of our review, we did note that the administration of two IM naloxone doses (0.8mg) has been insufficient in reversing all fentanyl overdoses, although the accuracy of such conclusions is contingent upon the quantity of fentanyl present in the drug samples consumed and the individual’s tolerance. For this reason community-based programs that solely distribute IM naloxone could pre-emptively begin distributing four or more doses to all program participants. Given that it is well established that overdose symptoms may recur after resuscitation, contingent upon the half-life of the specific opioid, keeping additional doses of naloxone on hand can be useful regardless of formulation.

Considering these findings and the current community practice of using multiple doses of standard IM and IN, we recommend providing at minimum, four standard doses of IN or IM naloxone to each individual, so that administration can continue until the recipient achieves stability, ensuring appropriate intervals between each dose, and extra doses are on hand in case of symptom recurrence. Given that some people who use fentanyl use multiple times per day, and some bystanders know multiple people who use fentanyl, providing an ample number of kits to potential bystanders is critical.

Higher-dosage formulations do not appear necessary for fentanyl overdoses and may in fact cause more harm than good as evidenced by the risk of precipitating opioid withdrawal. We did note that although there is little evidence that HDNF will save more lives, HDNF may elicit a faster overdose reversal rate and better outcomes for carfentanil overdoses if three or more doses of standard IN or IM are not available.

One barrier that remains in scaling up IM and IN naloxone is that only one brand of over-the-counter IN naloxone (Emergent) has been FDA approved. Approving generic naloxone and standard IM formulations will help speed up community-level naloxone coverage. Another barrier to carrying IM naloxone is that syringe possession remains illegal in some states.

### Data Limitations

Providing context to epidemiological and clinical data is important because it allows for a more accurate interpretation of the results. Without context, it is possible to make assumptions based on bias, or draw the wrong conclusions. This can lead to a lack of understanding of the underlying issues, and can result in ineffective or even harmful interventions. Additionally, it is also important to consider the potential biases that could influence the data, such as the socio-economic status, race, and gender of participants studied, that could impact the results and conclusions. Providing context to the data also allows for a better understanding of the limitations of the study and how generalizable the findings are. It also helps to ensure that the data is not misinterpreted or taken out of context. Additionally, much of the literature supporting the use of higher doses of naloxone fails to take into consideration the expressed needs, barriers, and consent of people who use drugs, which may have significant implications for the ethical and effective implementation of such interventions. For these reasons, we encourage scientists to speak to PWUD and service providers when developing and testing new HDNF products.

### Future Studies and Conclusion

More broadly, it is important to note that the majority of the research conducted in this area has not been done in settings that accurately reflect the contexts in which people who use drugs experience an overdose and withdrawal. More studies are needed to optimize community bystander reversals especially in the era of xylazine and other contaminants. Given that many people use alone, other interventions such as overdose prevention centers and overdose detection technologies may be necessary to better connect individuals to bystanders who are trained, equipped, willing and available to respond to potential overdoses.

In conclusion we did not find evidence to support HDNF. Community programs should provide at least four doses of standard IM or IN (and more if possible) to each program participant to optimize naloxone coverage without sacrificing the physical and psychological wellbeing of people who use drugs.

## List of Abbreviations

Abbreviation: Definition

FDA: Federal Drug Administration

IM: Intramuscular

IV: Intravenous

IN: Intranasal

HDNF: High-dose naloxone formulations

SUD: Substance use disorder(s)

PWUD: People who use drugs

EMS: Emergency Medical Services

SAMHSA: Substance Abuse and Mental Health Services Administration

CPR: cardiopulmonary resuscitation

MMWR: Morbidity and Mortality Weekly Report

PET: positron emission tomography

K_i_: Binding affinity

DPG: Daniel P. Garrett

PML: Paige M. Lemen

## Data Availability

All data produced in the present study are available upon reasonable request to the authors

https://tennesseeharmreduction.com/

## Acknowledgements

We thank the volunteers from Tennessee Harm Reduction and the Harm Reduction Innovation Lab who supported this work.

---

**Supplementary Table 1.**
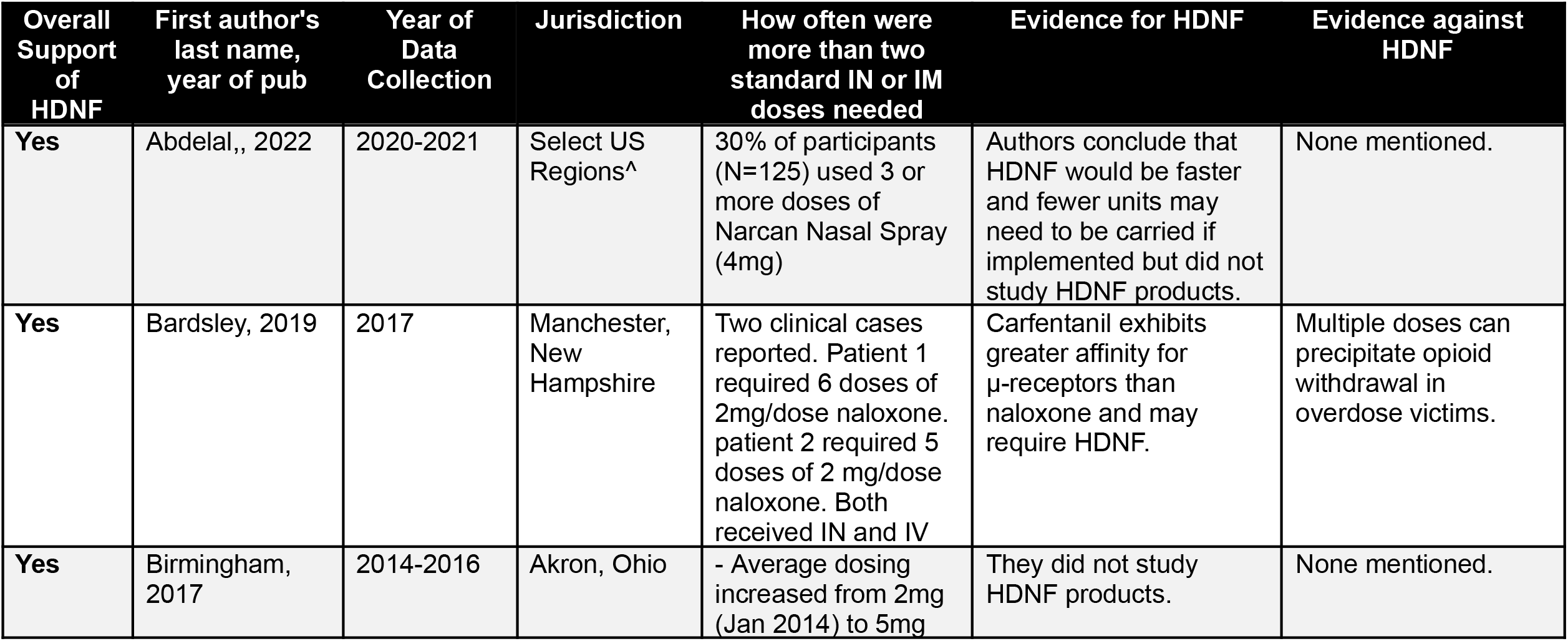
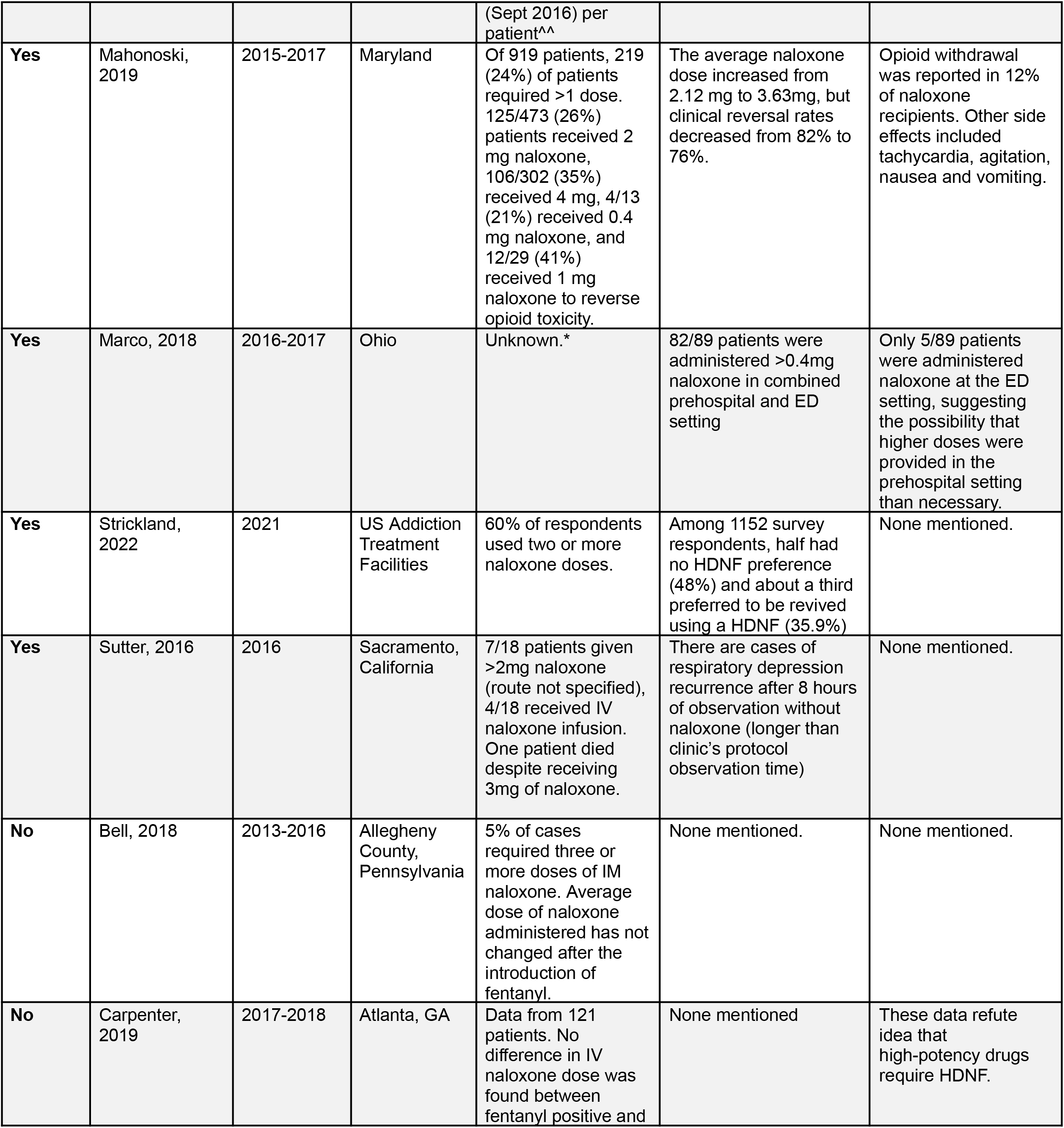
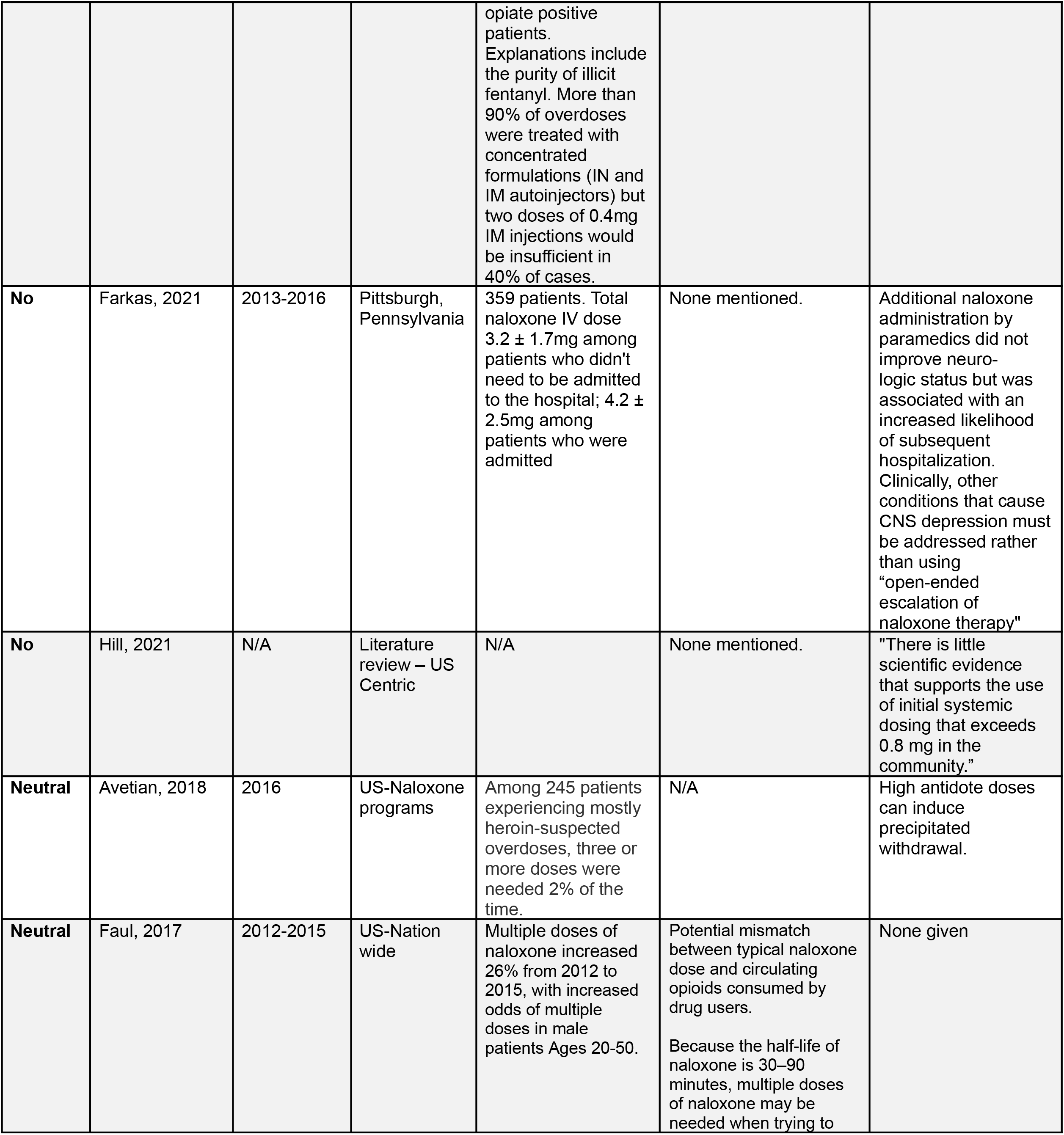
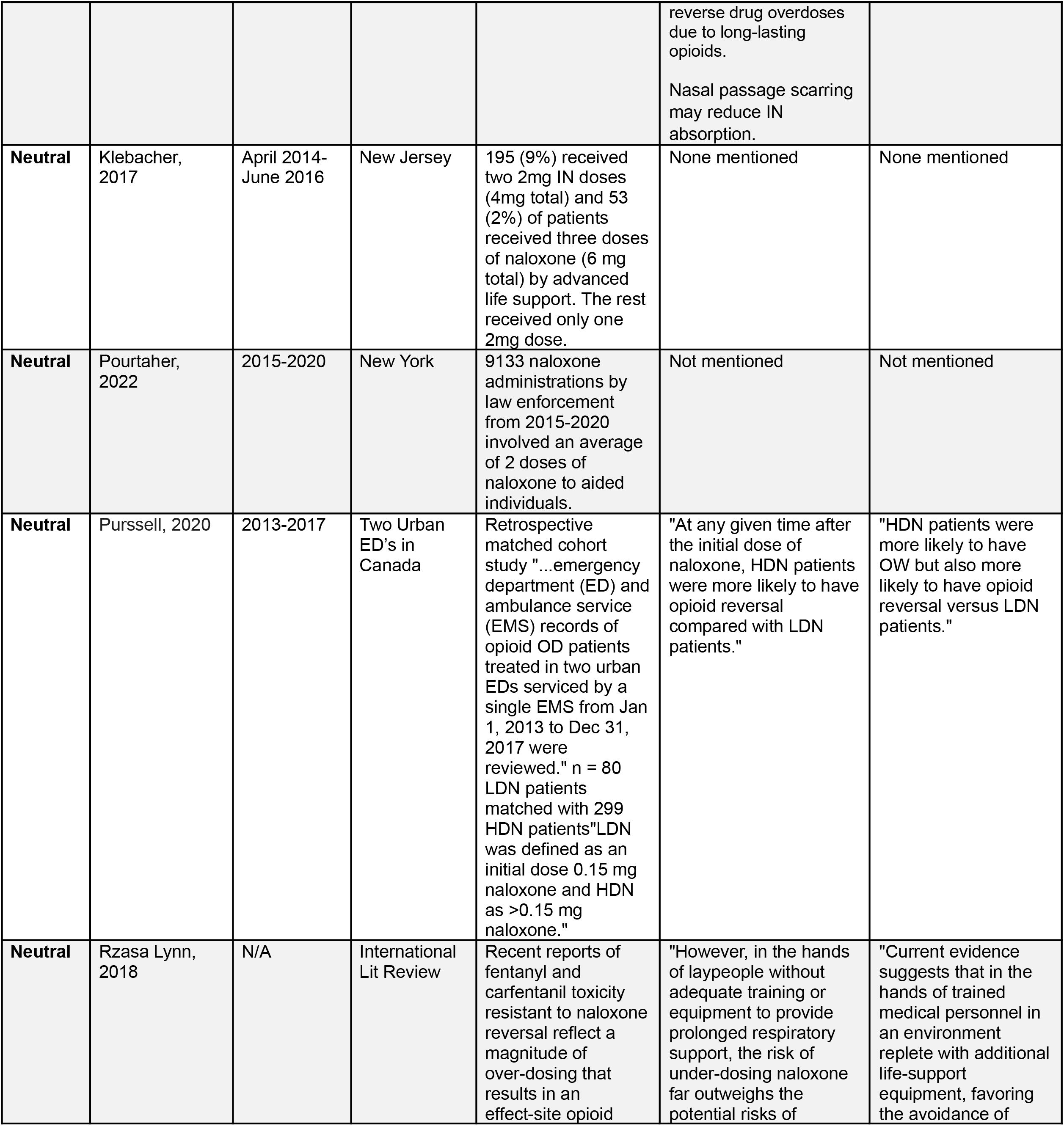
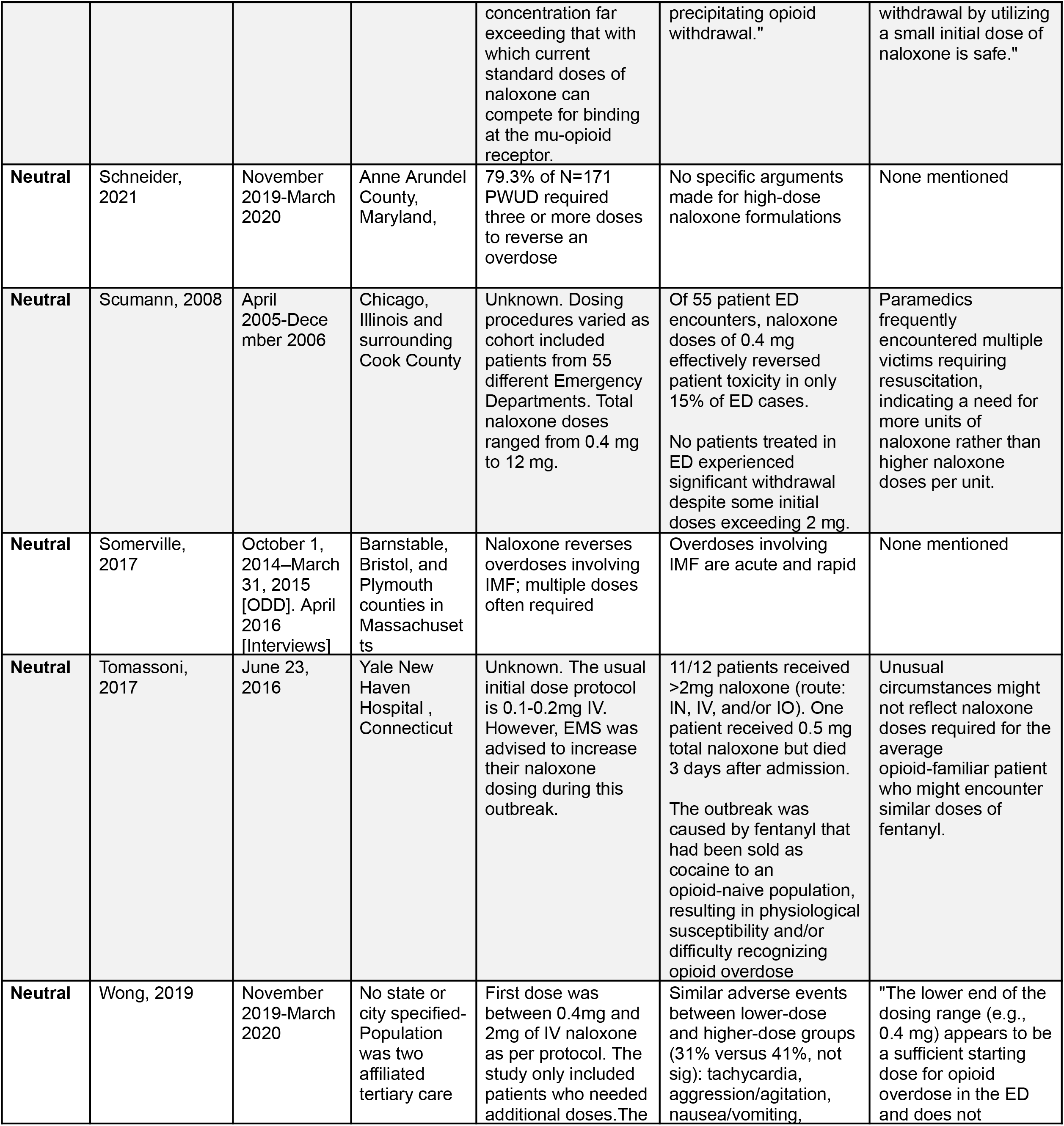
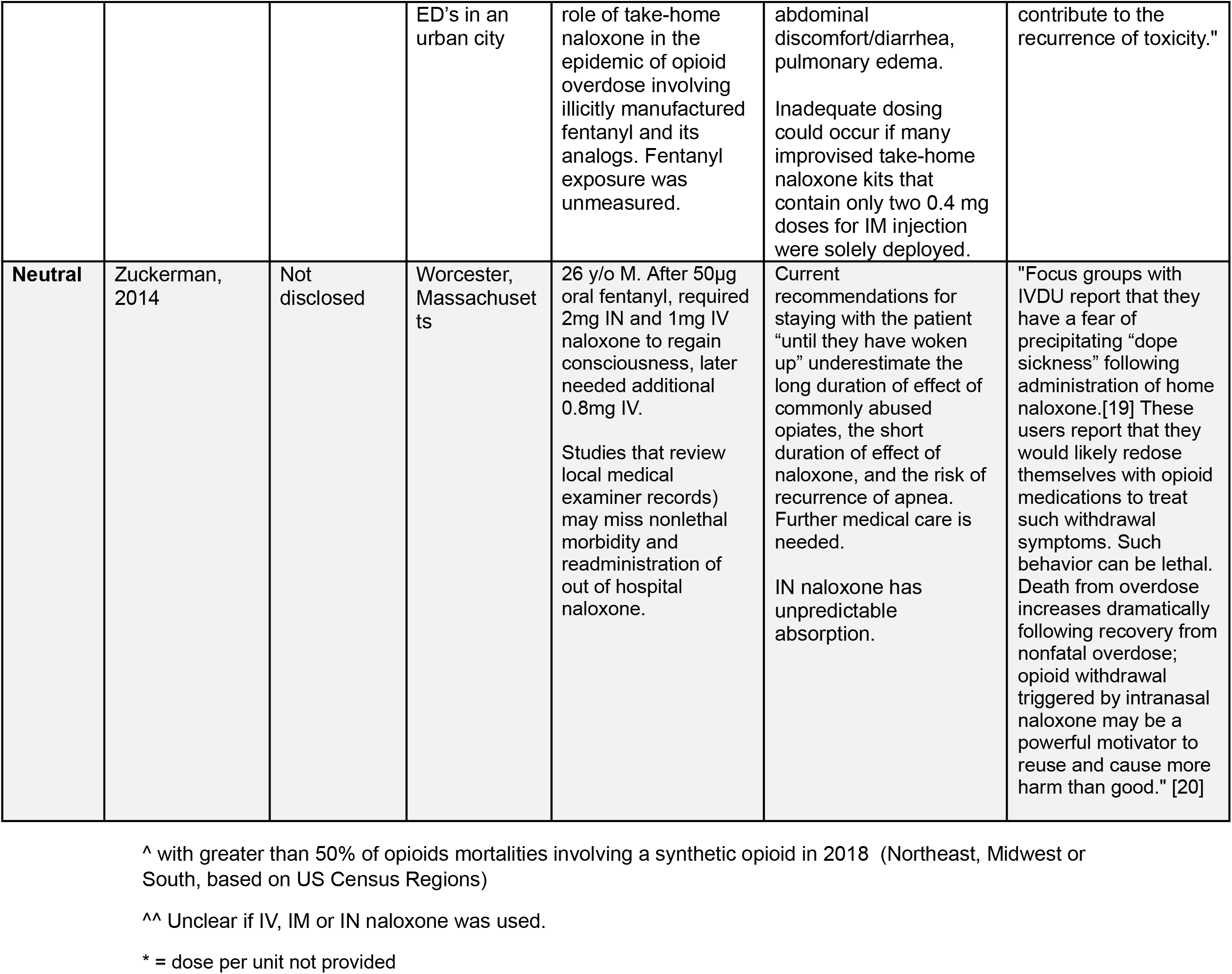
Published research articles involved in literature review.

